# Left common trunkus pulmonary veins have genetic background and poorer rhythm outcome after paroxysmal atrial fibrillation catheter ablation

**DOI:** 10.1101/2023.07.14.23292696

**Authors:** Sung Hwa Choi, Tae-Hoon Kim, Myunghee Hong, Oh-Seok Kwon, Daehoon Kim, Je-Wook Park, Hee Tae Yu, Jae-Sun Uhm, Boyoung Joung, Moon-Hyoung Lee, Chun Hwang, Hui-Nam Pak

## Abstract

**Background:** The genetic traits of pulmonary vein (PV) variants and rhythm outcomes after atrial fibrillation (AF) catheter ablation (AFCA) remain unclear. We explored the genetic and clinical characteristics and long-term rhythm outcomes of patients with AF and left common trunkus (LCT)-PVs or accessory PVs.

**Methods:** We included 2,829 patients with AF (74.0% men, age 59.1±10.7 years, 66.3% paroxysmal AF) and available genome-wide association study, cardiac computed tomography, and protocol-based regular rhythm follow-up results from the Yonsei AF ablation cohort database. We examined 1,223 single nucleotide polymorphisms in 12 genetic loci associated with AF and long-term rhythm outcomes after AFCA.

**Results:** We found LCT-PVs in 91(3.2%) and accessory PVs in 189(6.7%) patients. Rs9871453 (*SCN10A*) and rs1979409 (*NEO1*) were significantly associated with LCT-PV occurrence, and polygenic risk score (PRS) differed significantly between patients with LCT-PVs (p=1.64e-05) and normal PVs, but not those with accessory PVs (p=0.939). Patients with LCT-PVs had a higher proportion of the female sex(p=0.046) and CHA_2_DS_2_VASc score (p=0.026). After follow-up for 39.7±4.7 months, patients with LCT-PVs exhibited significantly greater LCT anterior wall thicknesses (p<0.001) and higher recurrence rate than those with normal PVs, particularly patients with paroxysmal AF (log-rank, p=0.042). LCT-PVs were independently associated with AF recurrence after AFCA (hazard ratio[HR], 2.26 [1.01–4.42]; p=0.046). Patients with LCT-PVs and higher PRSs had a higher risk of recurrent AF (adjusted HR 1.78, 95% CI 1.10–2.88, p=0.019).

**Conclusions:** Patients with LCT-PVs have a significant genetic background. Post-AFCA recurrence rate was significantly higher in patients with LCT-PVs and higher PRSs, particularly in those with paroxysmal AF.

**Clinical Perspective:** *What Is New?:* This study identifies specific genetic variants associated with the occurrence of LCT-PVs in AF patients undergoing catheter ablation. Higher AF recurrence rates were observed in LCT-PV patients, particularly those with paroxysmal AF. High-genetic risk LCT-PV patients exhibited increased AF recurrence and a thicker anterior wall of the left pulmonary vein compared to normal PV patients.

*What Are the Clinical Implications?:* The findings enhance our understanding of the genetic basis of AF and its anatomical manifestations, enabling personalized treatment approaches. Further research is needed to identify additional genetic variants associated with LCT-PV and to understand the recurrence of AF when using methods other than catheter ablation.

## Introduction

Atrial fibrillation (AF) is a common cardiac arrhythmia that affects millions of individuals worldwide, and its prevalence is increasing with the increasing age of the population. Although various risk factors for AF, including age, hypertension, diabetes, and structural heart disease, have been identified, recent studies have shown that genetic factors play an important role in the development of AF. Several variants of AF-associated genes, including *KCNQ1*, *NPPA*, *PITX2*, *SCN5a*, and *TBX5*, have been identified as risk factors for AF.^1-3^ These genes encode proteins involved in the regulation of ion channels, gap junctions, calcium handling, and atrial conduction, which are critical for the normal functioning of the heart. Certain pulmonary vein (PV) variations, such as the left common trunkus (LCT)-PV and accessory PV, are also associated with an increased risk of AF.^4^ However, the genetic traits of these PV variants and their impact on rhythm outcomes after AF catheter ablation (AFCA) remain unclear. AFCA is a common treatment for AF that involves ablating the PVs and other areas of the atrium associated with AF. Although AFCA can effectively restore sinus rhythm and improve symptoms, AF recurrence after the procedure remains a major clinical concern, particularly in patients with certain PV variations and genetic risk factors.

To address this knowledge gap, this study aimed to explore the genetic and clinical characteristics of patients with AF having an LCT-PV or accessory PV and their rhythm outcomes after AFCA. Specifically, we selected 1,223 single nucleotide polymorphisms (SNPs) from 12 AF-related genes to identify SNPs associated with LCT-PVs or accessory PVs. This study compared the calculated polygenic risk score (PRS) and PV antral wall thickness (WT) between patients with PV variations and those with normal PVs. It also investigated the relationship between PV variations, genetic risk factors, and AF recurrence after AFCA.

## Methods

### Study population

The study protocol was implemented in compliance with the principles of the Declaration of Helsinki and was approved by the Institutional Review Board of the Yonsei University Health System. All participants provided written informed consent for inclusion in the Yonsei AF ablation cohort study. From April 2009 to June 2021, of the patients included in the Precision Medicine Research Array (PMRA, Thermofisher Scientific, MA, USA) Yonsei Genome Wide Association Study (GWAS) AF ablation cohort, 2,829 patients were included in this study (Figure 1). This comprised 91 patients with LCT-PVs and 189 with accessory PVs. The exclusion criteria were as follows: 1) AF refractory to electrical cardioversion; 2) AF with rheumatic valvular disease; and 3) prior AF ablation or cardiac surgery. Each patient underwent computed tomography (CT) within 1 month before AFCA. We defined the LCT-PV as the separation between the upper and lower PVs >10 mm distal to the left PV antrum margin. The accessory PV was separate from the superior and inferior PVs, with independent drainage into the left atrium (LA). PV variations refer to cases with an LCT-PV or accessory PV.

**Figure 1.**
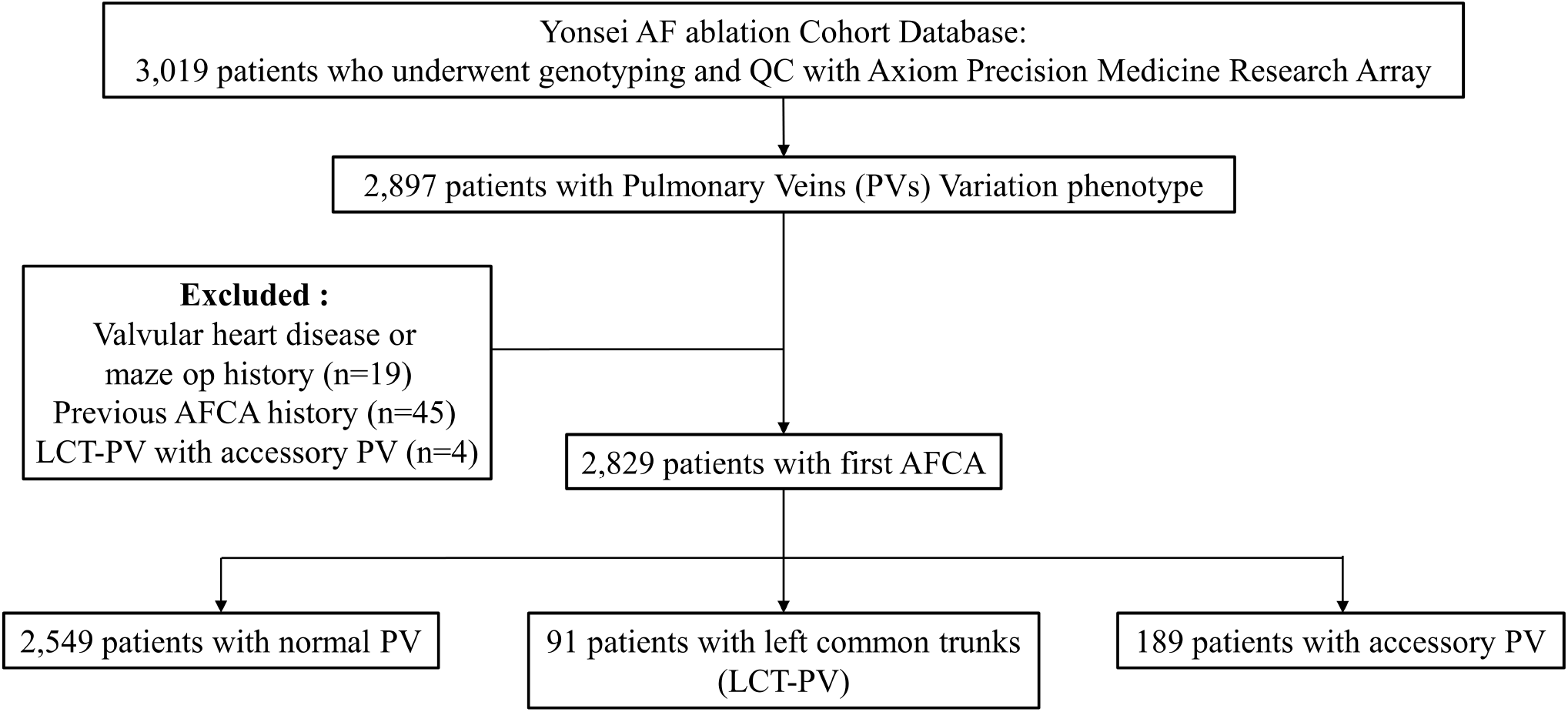
Flow chart of the study. AF, atrial fibrillation; AFCA, AF catheter ablation; LCT, left common trunkus; PV, pulmonary vein; QC, quality control

### Genotyping

Genomic DNA was extracted from peripheral blood samples using the QuickGene DNA Whole Blood Kit with QuickGene Mini 80 (KURABO, Osaka, Japan). DNA genotyping data were obtained using the Axiom PMRA. This DNA chip array consists of more than 900,000 markers to assist in the translation of research results to clinical insights.

### Selection of SNPs from 12 AF-associated genes

We examined 12 known AF-associated genes (*DSP, GJA1, HCN4, KCNQ1, NPPA, PITX2, RYR2, SCN5A, SHOX2, ATP2A2, TBX3, and TBX5;* Table S1) associated with the development of the heart and PVs.^5-8^ Initially, we selected 1,223 SNPs from these 12 candidate gene loci (within ± 250 kb). Subsequently, we divided the data into discovery and validation groups, based on the time of data collection. The discovery group consisted of patients recruited by October 2017 (n=1,550) and was used to identify SNPs associated with PV variants. SNPs identified in the discovery group were validated in a separate group of patients recruited after October 2017 (n=999). The validation group was used to confirm our findings and assess the reproducibility of the associations between the identified SNPs and PV variants. For further analysis, we identified three SNPs (rs146775048 and rs9871453 in the *SCN10A* locus, and rs1979409 in the *NEO1* locus) that showed a statistically significant association (p < 0.05) and consistent odds ratios (ORs) in both groups (Table 1). We defined a high PRS in patients with an LCT-PV by dividing the patients into two groups: those with no or one risk allele and those with two or more risk alleles, indicating a higher genetic risk. For patients with an accessory PV, we defined a high risk based on the presence or absence of the risk allele (rs146775048).

**Table 1.**
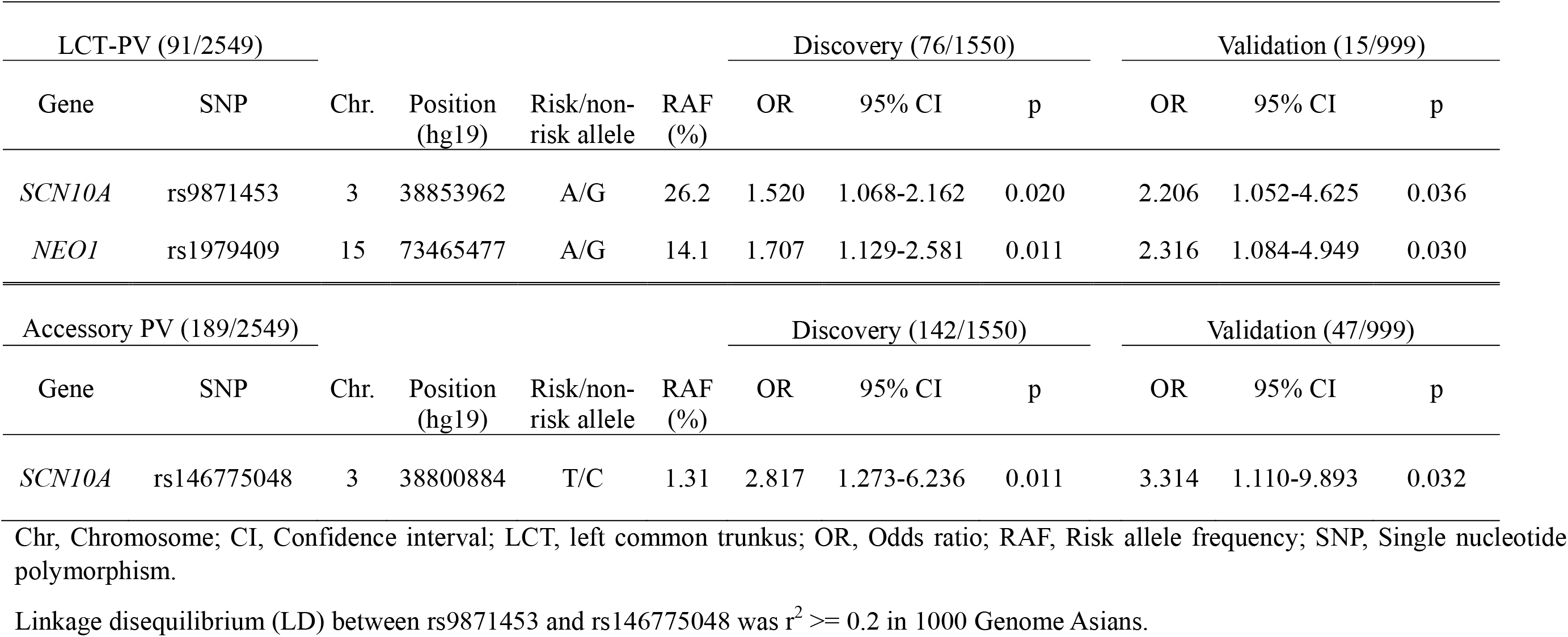
Association of LCT-PV and accessory PV for the *SCN10A* and *NEO1* SNPs.

### Evaluations of cardiac computed tomography and echocardiography

All patients underwent three-dimensional (3D) spiral CT using a 64 Channel Light Speed Volume CT scanner (Philips, Brilliance 63, Amsterdam, Netherlands). The CT scans were analyzed using an imaging processing workstation (Aquarius; Tera Recon, Inc., Foster City, CA, USA). The presence of normal PVs, LCT-PVs, or accessory PVs was determined using CT and 3D reconstruction of the images of the LA. Left atrial wall thickness (LAWT) and pericardial fat volume were measured using the AMBER software (Laonmed Inc., Seoul, Republic of Korea), as previously described.^9^

Transthoracic echocardiography was conducted twice: 3 months before the procedure and at the 1-year follow-up. The measurements included the dimensions of the LA, ejection fraction of the left ventricle, peak transmitral flow velocity (E), peak septal mitral annular velocity (Em) using tissue Doppler echocardiography, and other relevant parameters following the guidelines of the American Society of Echocardiography.

### Electrophysiological studies and catheter ablation

Electrocardiograms (ECGs) were recorded using a Prucka CardioLab^TM^ electrophysiology system (General Electric Medical Systems Inc., Milwaukee, WI, USA). We constructed 3D electroanatomical maps using the NavX (Abbott, Inc., Chicago, IL, USA) and CARTO (Biosense Webster, Diamond Bar, CA, USA) systems. The maps were created based on the location information obtained from a circumferential PV-mapping catheter (AFocus, Abbott, Inc., Chicago, IL, USA; Lasso, Biosense-Webster Inc., Diamond Bar, CA, USA) inserted through a long sheath. The 3D geometries of the LA and PVs were generated using a 3D mapping system and merged with the 3D spiral CT images. Multiview pulmonary venograms were acquired to ensure accurate alignment of the 3D map, CT scans, and fluoroscopy images of all patients. Contact bipolar electrograms were recorded from 500 to 1,000 points on the LA endocardium during high right atrial pacing at 500 ms, and the mean LA electrogram voltage was calculated.

For the AFCA procedure, we employed an open-irrigation catheter (Celsius, Johnson & Johnson Inc., Diamond Bar, CA, USA; NaviStar ThermoCool, Biosense Webster Inc., Diamond Bar, CA, USA; ThermoCool SF, Biosense Webster Inc.; ThermoCool SmartTouch, Biosense Webster Inc.; Coolflex, Abbott Inc., Minnetonka, MN, USA; 30–35 W; 47°C; FlexAbility, Abbott Inc.; ThermoCool SmartTouch, Biosense Webster Inc., and TactiCath, Abbott Inc.). All patients initially underwent circumferential PV isolation regardless of the PV anatomy. Additional linear or electrogram-guided ablation was performed in patients with AF at the discretion of the operator. After completing protocol-based ablation, AF or atrial tachycardia (AT) was induced using high-current burst pacing. An isoproterenol infusion was administered to increase the heart rate, and sustained AF or AT was treated with internal cardioversion using biphasic shock. To the best possible extent, AF triggers and atrial premature beats were ablated using quick 3D activation mapping.

Patients underwent regular follow-up visits at specified intervals after AFCA. These visits included ECG assessments and Holter monitoring according to established guidelines. AF recurrence was defined as any episode of AF or AT lasting at least 30 s, irrespective of anti-arrhythmic drug use. Clinical recurrence was determined by evidence of AF recurrence on ECG after a 3-month blanking period. Additional Holter or event-monitor recordings were obtained if patients reported palpitations.

### Statistical analysis

Descriptive statistics are reported as the mean ± standard deviation or median (interquartile range) for continuous variables and frequency and percentage for categorical variables.

Categorical variables were compared using the chi-square test, and continuous variables were compared using either the Wilcoxon rank-sum test or Student’s t-test, as appropriate. Logistic regression analysis was used to identify the risk factors for clinical recurrence and to estimate the ORs, 95% confidence intervals (CI), and p-values. Variables with p-values < 0.1, as determined by univariate analysis, were selected for multivariate analysis. Kaplan–Meier analysis with a log-rank test was used to analyze the probability of freedom from AF recurrence after AFCA. We calculated the PRS using the number of risk alleles. Statistical significance was set at p < 0.05. Statistical analyses were conducted using the R version 4.1.3 software (www.R-project.org; R Foundation for Statistical Computing, Vienna, Austria).

## Results

### Patient characteristics and genetic traits of PV variations

Of the 2,829 patients with AF (74.0% men, age 59.1±10.7 years, 66.3% paroxysmal AF) with available genome-wide association study results, 91 (3.2%) patients had an LCT-PV and 189 (6.7%) had an accessory PV. Table 2 summarizes the patient characteristics comparing those with LCT-PVs and accessory PVs. Patients with LCT-PVs had a higher proportion of the female sex (p=0.046), higher E/Em ratios (p=0.002), and lower LA voltages (p=0.007). They were older (p=0.025), had higher CHA_2_DS_2_-VASc scores (p=0.026) at the time of ablation, and more frequently underwent empirical extra-PV LA ablation (p=0.012).

**Table 2.**
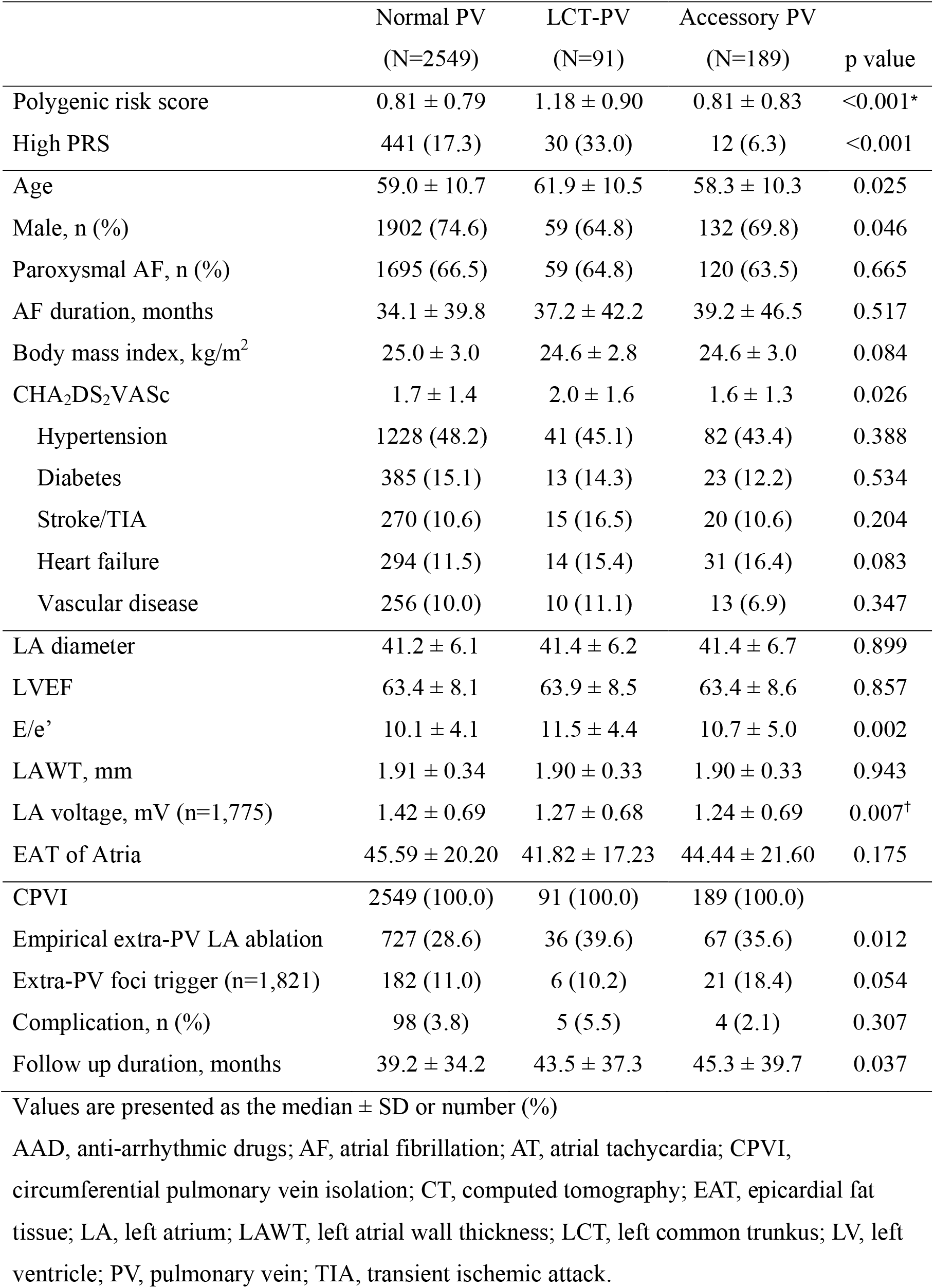
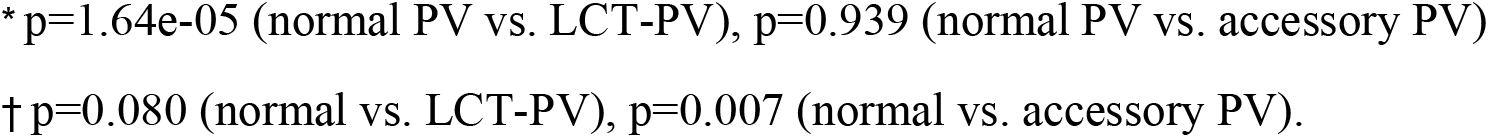
Baseline characteristics according to LCT-PV and accessory PV.

|  | Normal PV<br>(N=2549) | LCT-PV<br>(N=91) | Accessory PV<br>(N=189) | p value |
| --- | --- | --- | --- | --- |
| Polygenic risk score | 0.81 ± 0.79 | 1.18 ± 0.90 | 0.81 ± 0.83 | <0.001* |
| High PRS | 441 (17.3) | 30 (33.0) | 12 (6.3) | <0.001 |
| Age | 59.0 ± 10.7 | 61.9 ± 10.5 | 58.3 ± 10.3 | 0.025 |
| Male, n (%) | 1902 (74.6) | 59 (64.8) | 132 (69.8) | 0.046 |
| Paroxysmal AF, n (%) | 1695 (66.5) | 59 (64.8) | 120 (63.5) | 0.665 |
| AF duration, months | 34.1 ± 39.8 | 37.2 ± 42.2 | 39.2 ± 46.5 | 0.517 |
| Body mass index, kg/m<sup>2</sup> | 25.0 ± 3.0 | 24.6 ± 2.8 | 24.6 ± 3.0 | 0.084 |
| CHA<sub>2</sub>DS<sub>2</sub>VASc | 1.7 ± 1.4 | 2.0 ± 1.6 | 1.6 ± 1.3 | 0.026 |
| Hypertension | 1228 (48.2) | 41 (45.1) | 82 (43.4) | 0.388 |
| Diabetes | 385 (15.1) | 13 (14.3) | 23 (12.2) | 0.534 |
| Stroke/TIA | 270 (10.6) | 15 (16.5) | 20 (10.6) | 0.204 |
| Heart failure | 294 (11.5) | 14 (15.4) | 31 (16.4) | 0.083 |
| Vascular disease | 256 (10.0) | 10 (11.1) | 13 (6.9) | 0.347 |
| LA diameter | 41.2 ± 6.1 | 41.4 ± 6.2 | 41.4 ± 6.7 | 0.899 |
| LVEF | 63.4 ± 8.1 | 63.9 ± 8.5 | 63.4 ± 8.6 | 0.857 |
| E/e' | 10.1 ± 4.1 | 11.5 ± 4.4 | 10.7 ± 5.0 | 0.002 |
| LAWT, mm | 1.91 ± 0.34 | 1.90 ± 0.33 | 1.90 ± 0.33 | 0.943 |
| LA voltage, mV (n=1,775) | 1.42 ± 0.69 | 1.27 ± 0.68 | 1.24 ± 0.69 | 0.007<sup>†</sup> |
| EAT of Atria | 45.59 ± 20.20 | 41.82 ± 17.23 | 44.44 ± 21.60 | 0.175 |
| CPVI | 2549 (100.0) | 91 (100.0) | 189 (100.0) |  |
| Empirical extra-PV LA ablation | 727 (28.6) | 36 (39.6) | 67 (35.6) | 0.012 |
| Extra-PV foci trigger (n=1,821) | 182 (11.0) | 6 (10.2) | 21 (18.4) | 0.054 |
| Complication, n (%) | 98 (3.8) | 5 (5.5) | 4 (2.1) | 0.307 |
| Follow up duration, months | 39.2 ± 34.2 | 43.5 ± 37.3 | 45.3 ± 39.7 | 0.037 |

Values are presented as the median ± SD or number (%)
AAD, anti-arrhythmic drugs; AF, atrial fibrillation; AT, atrial tachycardia; CPVI, circumferential pulmonary vein isolation; CT, computed tomography; EAT, epicardial fat tissue; LA, left atrium; LAWT, left atrial wall thickness; LCT, left common trunkus; LV, left ventricle; PV, pulmonary vein; TIA, transient ischemic attack.
\* $p=1.64e-05$ (normal PV vs. LCT-PV), $p=0.939$ (normal PV vs. accessory PV)
† $p=0.080$ (normal vs. LCT-PV), $p=0.007$ (normal vs. accessory PV).

Two SNPs (rs9871453 in the *SCN10A* locus and rs1979409 in the *NEO1* locus) were associated with LCT-PVs (Table 1), and the PRS calculated based on these SNPs was significantly higher than that for normal PVs (p=1.64e-05, Figure 2A). In contrast, one SNP (rs146775048 in the *SCN10A* locus) was associated with accessory PVs, but the PRS based on this SNP did not differ significantly from that for normal PVs (p=0.939). The high-genetic-risk group, defined by the presence of risk alleles, constituted 33% of patients with LCT-PVs, which was significantly higher than that of patients with normal PVs (17.3%), while accounting for 6.3% of patients with accessory PVs, which was lower than that of normal patients (p<0.001, Table 2).

**Figure 2.**
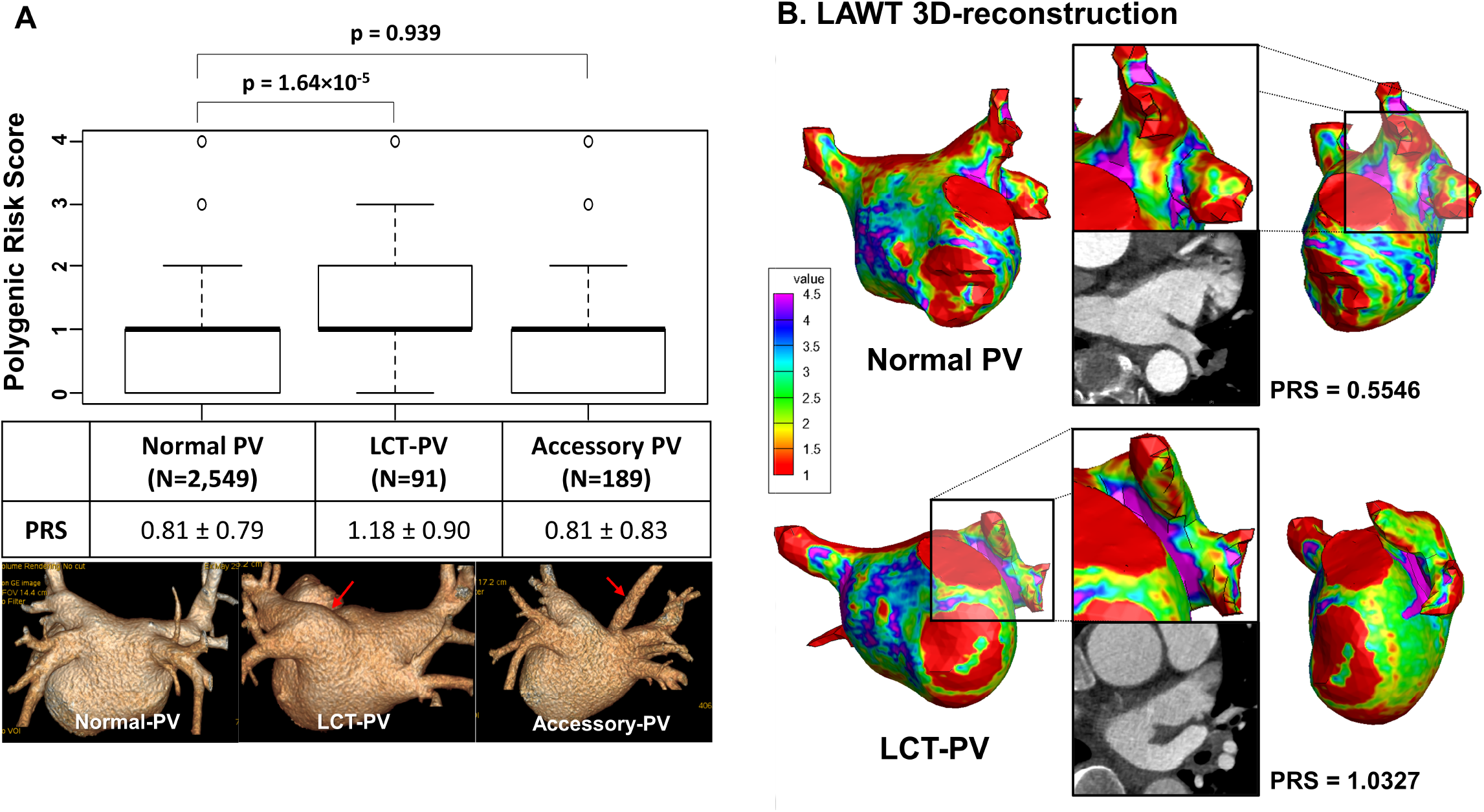
Comparison of PRS and the representative images of the LCT-PV and accessory PV (A) LAWT 3D-reconstruction CT images of a patient with LCT-PV and a patient with normal PV (B) PRS calculated based on associated SNPs was significantly higher in patients with LCT-PV than in normal PV. However, in patients with accessory PV, there was no significant difference from normal PV. The legend on the left indicates the left atrial wall thickness (mm) by color. The left anterior PV thickness in the LCT-PV group, shown in purple, was larger than normal PV. AF, atrial fibrillation; CT, Computed tomography; LAWT, left atrial wall thickness; LCT, left common trunkus; PRS, Polygenic risk score; PV, pulmonary vein

### AFCA outcomes in patients with LCT-PVs

During the follow-up period of 39.7 ± 34.8 months, the overall clinical recurrence rates did not differ among patients with LCT-PVs, accessory PVs, and normal PVs (log-rank p=0.510, Figure 3A). Patients with LCV-PVs showed significantly higher clinical recurrence rates than patients with paroxysmal AF (log-rank p=0.042, Figure 3B), but the clinical recurrence rates were not significantly higher than those with persistent AF (log-rank p=0.580, Figure 3C). Comparisons of patients paroxysmal AF with LCT-PVs and normal PVs are shown in Table S2.

**Figure 3.**
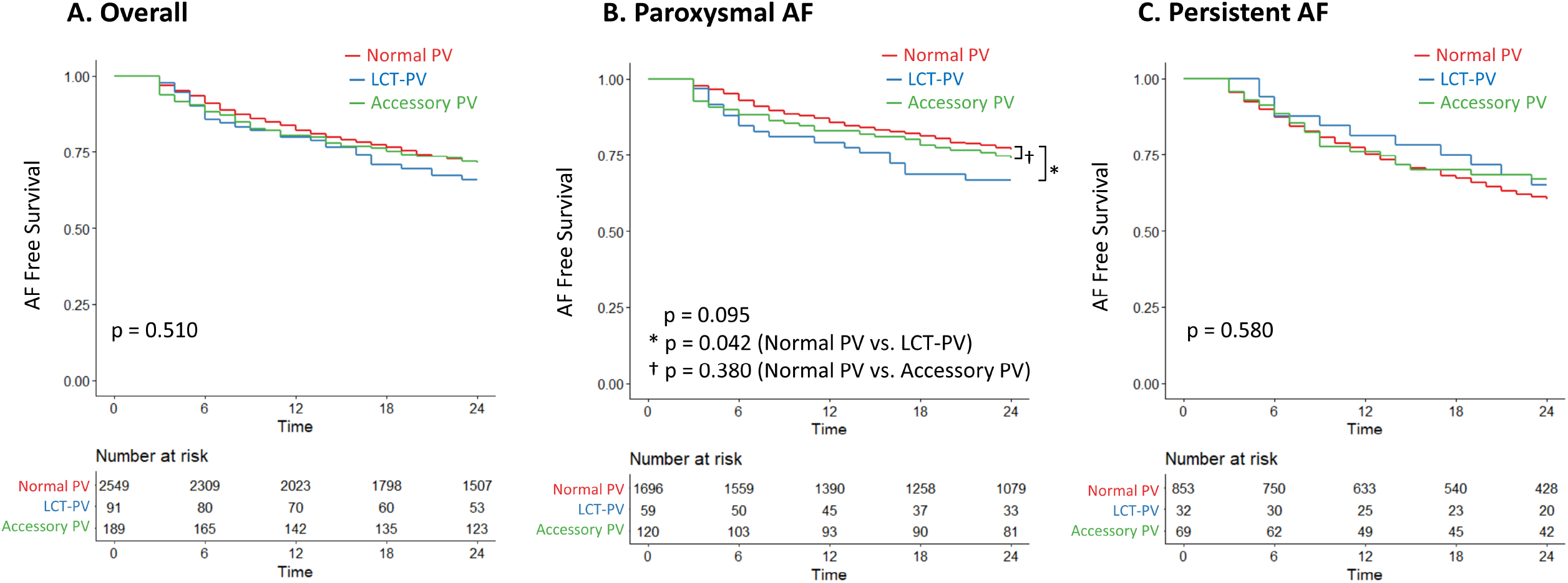
Kaplan–Meier curves of AF relapse after AFCA categorized by the types of AF. Patients with LCV-PV showed a significantly higher clinical recurrence rate in patients with paroxysmal AF, but did not have a significantly higher clinical recurrence rate in patients with persistent AF. AF, atrial fibrillation; AFCA, AF catheter ablation; LCT, left common trunkus; PV, pulmonary vein;

A multivariate logistic regression analysis revealed that clinical recurrence was independently associated with the presence of LCT-PVs (hazard ratio [HR] 2.26 [1.01–4.42], p=0.046), higher early recurrence (HR 2.02 [1.47–2.79], p<0.001), lower LA voltage (HR 0.64 [0.48, 0.85], p=0.002), and higher extra PV foci triggers (HR 1.71 [1.18, 2.50], p=0.005), but not accessory PVs (Table 3).

**Table 3.** Predictors of AF recurrence on univariate and multivariate Cox regression analysis.

|  | Univariate |  | Multivariate |  |
| --- | --- | --- | --- | --- |
|  | HR (95% C.I.) | *P* value | HR (95% C.I.) | *P* value |
| Age, years | 1.00 [1.00, 1.01] | 0.250 | 0.98 [0.97, 1.00] | 0.098 |
| Male, n(%) | 0.96 [0.85, 1.10] | 0.556 | 0.87 [0.61, 1.25] | 0.458 |
| Paroxysmal AF, n(%) | 0.56 [0.50, 0.63] | <0.001 | 0.91 [0.64, 1.30] | 0.602 |
| AF duration, months | 1.00 [1.00, 1.00] | 0.020 | 1.00 [1.00, 1.00] | 0.432 |
| Body mass index, kg/m<sup>2</sup> | 1.02 [1.00, 1.04] | 0.118 |  |  |
| Hypertension | 1.17 [1.04, 1.31] | 0.009 | 1.15 [0.84, 1.56] | 0.386 |
| Diabetes | 1.02 [0.87, 1.20] | 0.780 |  |  |
| Stroke/TIA | 1.24 [1.04, 1.47] | 0.019 | 1.24 [0.76, 2.02] | 0.381 |
| Heart failure | 1.33 [1.12, 1.57] | 0.001 | 1.06 [0.67, 1.67] | 0.806 |
| Vascular disease | 0.97 [0.81, 1.17] | 0.761 |  |  |
| Early recurrence | 2.80 [2.49, 3.15] | <0.001 | 2.02 [1.47, 2.79] | <0.001 |
| Presence of LCT-PV | 1.30 [0.96, 1.76] | 0.093 | 2.12 [1.01, 4.42] | 0.046 |
| Presence of accessory PV | 1.04 [0.84, 1.30] | 0.704 |  |  |
| rs146775048 (*SCN10A*) | 1.17 [0.82, 1.67] | 0.399 |  |  |
| rs9871453 (*SCN10A*) | 1.06 [0.97, 1.17] | 0.211 |  |  |
| rs1979409 (*NEO1*) | 1.03 [0.91, 1.15] | 0.682 |  |  |
| Polygenic risk score | 1.06 [0.98, 1.14] | 0.140 |  |  |
| LA diameter (Echo) | 1.04 [1.03, 1.05] | <0.001 | 1.02 [0.99, 1.05] | 0.230 |
| LV ejection fraction, % | 0.99 [0.98, 0.99] | <0.001 | 0.99 [0.97, 1.01] | 0.454 |
| E/e' | 1.01 [1.00, 1.03] | 0.051 | 0.96 [0.91, 1.02] | 0.203 |
| LA wall stress | 1.00 [1.00, 1.00] | <0.001 | 1.00 [1.00, 1.00] | 0.788 |
| EAT volume of atria (cm<sup>3</sup>, CT) | 1.01 [1.00, 1.01] | <0.001 | 1.00 [0.99, 1.01] | 0.501 |
| LAWT, mean | 1.05 [0.88-1.25] | 0.605 |  |  |
| Mean LA voltage, mV (n=1,775) | 0.60 [0.53, 0.68] | <0.001 | 0.64 [0.48, 0.85] | 0.002 |
| Extra-PV foci trigger (n=1,824) | 1.87 [1.52, 2.28] | <0.001 | 1.71 [1.18, 2.50] | 0.005 |

AF, atrial fibrillation; C.I., confidence interval; CT, computed tomography; EAT, epicardial
adipose tissue; LA, left atrium; LAWТ, left atrial wall thickness; LCT, left common trunkus;
LV, left ventricle; HR, hazard ratio; PV, pulmonary vein; TIA, transient ischemic attack.

### Prognostic value of the PRS and LAWT in patients with LCT-PVs

Patients with LCT-PVs having a high PRS exhibited significantly higher rates of AF recurrence than others (log-rank p=0.025, Figure 4A) and were independently associated with clinical recurrence of AF after AFCA (adjusted HR 1.78 [1.10–2.88], p=0.019, Table S3, Figure 4C). However, the presence of accessory PVs with a high PRS did not affect rhythm outcomes after AFCA (log-rank p=0.480, Figure 4B).

**Figure 4.**
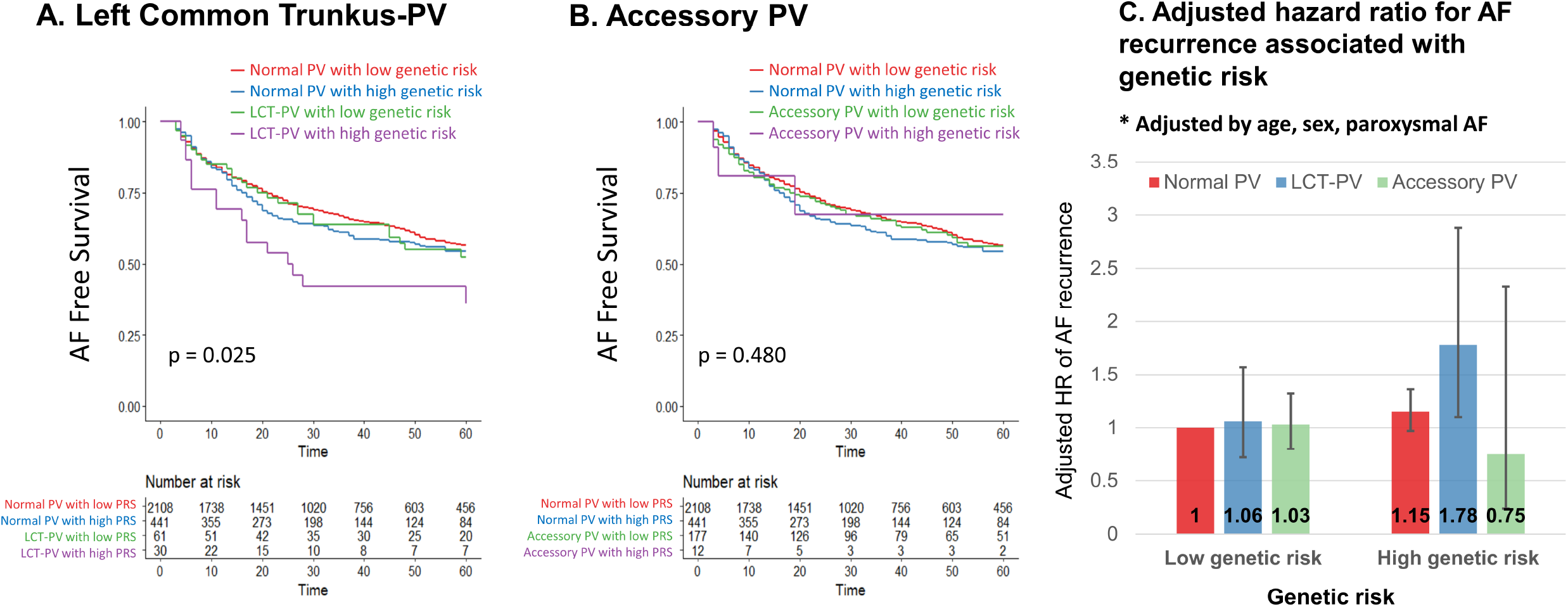
Kaplan–Meier curves of freedom from recurrence of atrial fibrillation after AFCA according to presence of LCT-PV with genetic risk (A) and the accessory PV with genetic risk (B), adjusted hazard ratio for AF recurrence associated with genetic risk (C) Patients with LCT-PVs having a high PRS exhibited significantly higher rates of AF recurrence than others. However, the presence of accessory PVs with a high PRS did not affect rhythm outcomes after AFCA AF, atrial fibrillation; AFCA, AF catheter ablation; LCT, left common trunkus; PV, pulmonary vein

In the analyses of LAWT, anterior left PV thickness in the LCT-PV group was greater than that in the normal or accessory PV group (3.57 ± 1.33 mm in the LCT-PV, 3.06 ± 0.88 mm in the normal PV, and 3.17 ± 0.93 mm in the accessory PV groups, p<0.001, Table S4), while the posterior left PV thickness in the LCT-PV group was lesser than that of the other groups (1.38 ± 0.39 mm in the LCT-PV, 1.64 ± 0.40 mm in the normal PV, and 1.63 ± 0.36 mm in the accessory PV groups, p<0.001). The anterior WT of the left PV was greater in patients with LCT-PVs having a high PRS (p<0.001) than in those with LCT-PVs having a low PRS or in other groups (Table 4). A representative image comparing 3D-reconstruction of the CT images of a patient with an LCT-PV and a patient with a normal PV with an LWAT map is presented in Figure 2B.

**Table 4.**
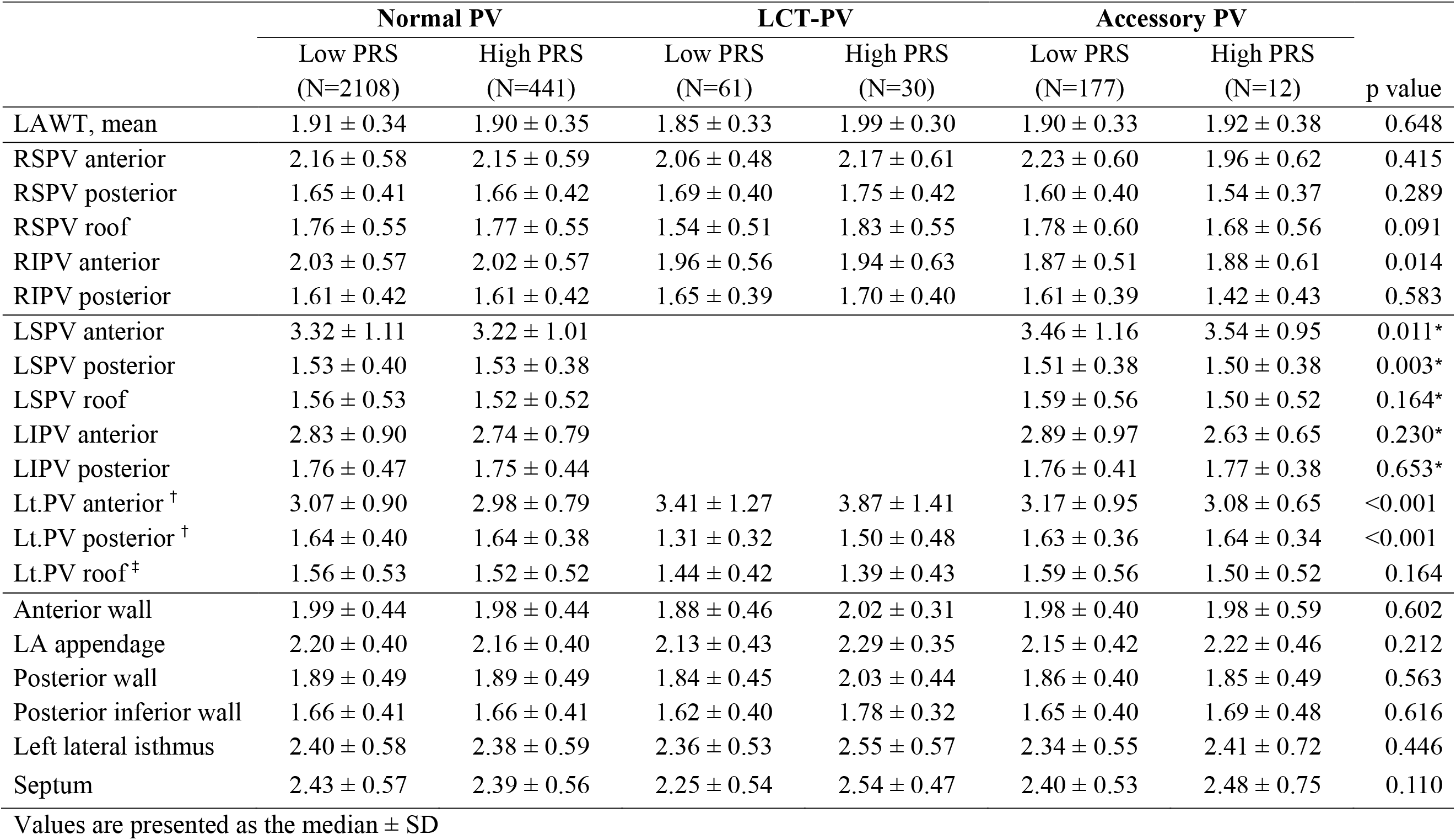
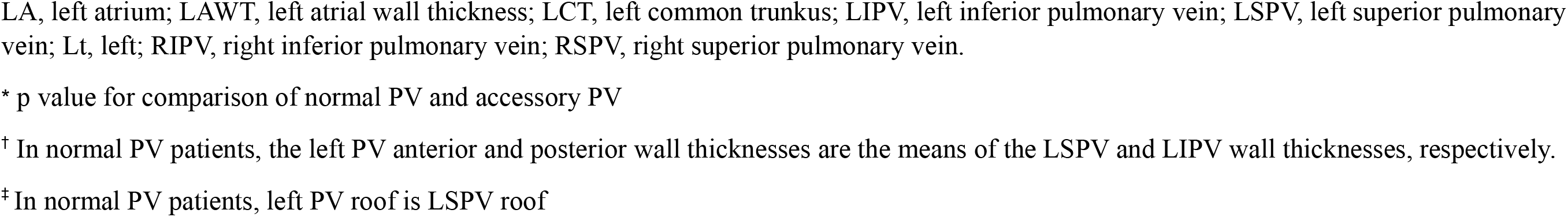
LAWT according to PV variants with genetic risk.

## Discussion

### Main findings

In this study, we analyzed the genetic associations and characteristics of PV variations and outcomes after AFCA using a retrospective cohort database with available GWAS data from a single institution. By analyzing a database of 2,829 patients, we identified associations between specific SNPs and the presence of PV variants. Patients with LCT-PVs exhibited a significantly higher recurrence rate than those with paroxysmal AF. The presence of LCT-PVs, higher early recurrence rates, lower LA voltages, and additional PV foci triggers were independently associated with clinical recurrence. Furthermore, patients with an LCT-PV and a high PRS had a significantly higher rate of recurrent AF of 78% than patients with a normal PV and low PRS. Patients with an LCT-PV showed a greater anterior WT of the left PV than those with a PV and an accessory PV. Moreover, in patients with LCT-PVs, those with a high PRS exhibited a greater anterior WT of the left PV than those with a low PRS.

### Incidence of PV variations and their effects on AF

The exact prevalence of anatomical variants of PVs is unknown because of insufficient epidemiological data.^10^ The occurrence of LCT-PVs is common with an incidence of 12-37%.^11,12^ Accessory PVs, which occur in 6-20% of the population, are common on the right side, and the right middle accessory vein is most frequently observed. The effect of the occurrence of LCT-PVs on AF rhythm control is unknown. PV variations have been reported in approximately 25% of patients with AF, and several studies have shown a higher prevalence of LCT-PVs in patients with AF than in controls.^4,13,14^ Therefore, it can be speculated that LCT-PVs will have some influence in the development of AF.

### Impact of characteristics of LCT-PVs on the outcome of ablation

The effect of LCT-PVs on PV isolation during AFCA and variations in outcomes depending on the procedure are unknown.^15^ Although a previous study reported that the presence of an LCT-PV did not affect outcomes in patients with paroxysmal AF undergoing cryoablation, few patients with LCT-PVs were included in that study (17.5%, n=14).^16^ Further, it showed that the anatomical indicators of the PV area and PV ovality did not predict AF recurrence. However, another study reported that the presence of LCT-PVs was independently associated with AF recurrence in patients with paroxysmal AF who underwent AFCA.^12^ In this study, recurrence after AFCA was high in patients with LCT-PVs and paroxysmal AF, confirming that the presence of LCT-PVs was independently associated with AF recurrence.

A previous study reported that the left intervenous ridge length affected AF recurrence in patients with LCT-PVs.^11^ The thicker and more complex arrangement of myocardial fibers in the left intervenous ridge makes PV isolation more difficult in the presence of more extensive ridges. In one study, PV reconnection was confirmed during repeat AFCA in most patients with LCT-PVs. In particular, there was a connection with the LCT-PV inferior site.^17^ Our study showed a difference in the thickness of the left PV, especially in the anterior wall of the LCT-PV. A thicker anterior wall may make complete isolation of the LCT-PV from the LA difficult, which is a critical step in AF ablation. However, the impact of anterior WT on AF ablation outcomes can vary depending on several factors, such as the location and extent of ablation, ablation technique used, and operator experience.^18^ In some cases, thicker anterior walls may be successfully ablated using appropriate techniques and equipment and therefore, may not necessarily lead to poor outcomes.^19^

In contrast, when comparing patients with a normal anatomy and those with an LCT-PV, the left atrial diameter, total average left atrial WT, or epicardial fat tissue did not differ significantly, but LA voltage and E/e’ differed significantly. A low LA voltage is associated with increased susceptibility to AF and poor outcomes after catheter ablation. This may be because a low LA voltage reflects underlying structural changes in the atrial myocardium, such as fibrosis, which can create a proarrhythmic substrate for AF.^20^ In addition, an elevated LA pressure predicted by E/e’ is also associated with electrophysiological LA remodeling and contributes to AF recurrence.^21^

### Impact of PRS of the LCT-PV on the outcome of ablation

The PRS is calculated based on a combination of multiple genetic variants associated with a particular trait or disease, such as AF.^3^ In our study, we identified several genetic variants associated with risk of LCT-PV formation. In addition to its potential use in predicting the LCT-PV anatomy, an LCT-PV PRS could also be used to identify individuals at an increased risk of AF, as the occurrence of an LCT-PV is a known risk factor for AF. In patients with AF, a higher AF-PRS is associated with a more advanced LA electrical substrate, with regions of increased conduction velocity heterogeneity, slowed conduction, complex fractionated signals, and bipolar voltage heterogeneity.^22^ Falasconi et al. reported that during the initial procedure, the absence of first-pass and reconnection sites were identified during repeat procedures to be predominantly located in the thickest regions of the PV antrum.^23^ According to the results of that study, the use of LAWT-guided PV antrum isolation demonstrated favorable outcomes. We found that in patients with a high PRS, a thicker anterior WT of the left PV was confirmed, and AF recurrence occurred more frequently after ablation. These results suggest that the LCT-PV PRS can potentially be used as a predictive marker to identify individuals who may benefit from a tailored procedure for AF.

### Clinical implications

Understanding the impact of PV variations on the outcomes of ablation procedures is important because it can help clinicians make informed decisions regarding the most effective treatment options for patients with AF. PV anatomical variations can affect the success of the procedure, and identifying these variations before the procedure can help clinicians plan a more individualized approach to ablation, improving the chances of success and reducing the risk of complications.

Knowledge of the genetic associations and characteristics of PV variants can provide important insights into mechanisms underlying the development of AF. In addition, a PRS associated with LCT-PV formation could potentially be used to identify individuals at an increased risk of LCT-PV formation and guide clinical decision-making, including preprocedural evaluation and risk stratification of patients with AF.

### Limitations

This study had some limitations. First, the study was conducted at a single center, which may limit the generalizability of the findings to other populations. Second, the study only included patients who underwent catheter ablation; therefore, the results may not be applicable to patients treated using other ablation methods, such as cryoablation or pulse field ablation. Lastly, the clinical utility of PRS remains limited, as genetic testing is not routinely performed in patients with AF, and the role of genetic risk assessment has not been fully established in the current guidelines. Further research is needed to validate the clinical utility of the LCT-PV PRS and to determine its optimal use in clinical practice.

## Conclusions

PRS calculated using two associated SNPs was significantly higher in patients with LCT-PVs than in those with normal PVs. Patients with LCT-PVs exhibited a higher recurrence rate, especially those with paroxysmal AF, and had a significantly greater left PV anterior WT than other groups. Additionally, patients with an LCT-PV and a high PRS had a higher risk of recurrent AF. These findings suggest that understanding the impact of PV variations on AFCA outcomes, including genetic factors, could help develop personalized treatment approaches for patients with AF.

## Nonstandard Abbreviations and Acronyms

AAD: Anti-arrhythmic drug
AF: Atrial fibrillation
AFCA: AF catheter ablation
CPVI: Circumferential pulmonary vein isolation
CT: Computed tomography
ECG: Electrocardiography/electrocardiogram
LA: Left atrium/atrial
LAVI: Left atrial volume index
LCT: left common trunkus
LV: Left ventricle/ventricular
PV: Pulmonary vein
PRS: Polygenic risk score

## Acknowledgments

**Sources of Funding**

This work was supported by a grant [HI21C0011] from the Ministry of Health and Welfare and a Korea Medical Device Development Fund grant [Project number 1711174471; RS-2022-00141473] funded by the Ministry of Science and ICT, Ministry of Trade, Industry, and Energy, Ministry of Health & Welfare, and Ministry of Food and Drug Safety of the Korean government.

## Data availability

The data underlying this article will be shared on reasonable request to the corresponding author.

